# Development of a protocol for soil transmitted helminths DNA extraction from feces by combining commercially available solutions

**DOI:** 10.1101/2022.12.12.22283332

**Authors:** AA Devyatov, EE Davydova, AR Luparev, SA Karseka, AK Shuryaeva, AV Zagainova, GA Shipulin

## Abstract

**Background:** One of the main challenges for the mass introduction of molecular diagnostics of soil-transmitted helminths (STHs) into clinical practice is the lack of a generally recognized effective method for isolating parasitic DNA from fecal samples. In the present study, we assessed the effect of various pretreatment procedures on the efficiency of removing PCR inhibitors and extracting *Toxocara canis* DNA from feces.

**Methodology and main results:** In the first part of the work, we evaluated the effectiveness of four destructive methods (bead beating, the action of temperature-dependent enzymes, freeze-heat cycles, and incubation in a lysis buffer of a commercial kit) on the integrity of Toxocara eggs using microscopy and the efficiency of DNA extraction using PCR. Our results showed that Toxocara eggs were most effectively destroyed using the bead beating procedure, while the effect of enzymes and freeze-heat cycles did not lead to significant destruction of the eggs or the release of Toxocara DNA. In the second part of the work, we evaluated the effect of prewashes with 0.1% Tween-20 solution and the use of commercial concentrators on DNA extraction from fecal samples contaminated with *T. canis* eggs. We have shown that the use of commercial concentrators in combination with sample washing can significantly increase the DNA yield and reduce PCR inhibition.

**Conclusions:** A bead beating procedure for 30 minutes at a shaking frequency of 50 Hz was sufficient to completely destroy the *Toxocara canis* eggs. Helminth DNA isolation protocols that do not include a bead beating step are not preferred. The use of a commercial concentrator followed by washing with a 0.1% Tween-20 solution can significantly increase the yield of STHs DNA and reduce PCR inhibition.

**AUTHOR SUMMARY:** DNA-based techniques are increasingly being used for the diagnosis of intestinal helminth infections in both clinical and research laboratories. However, extracting DNA from intestinal worm eggs in feces remains a challenge because the very tough eggshell protects their DNA. In addition, feces contain inhibitors that can interfere with test results, and these must be removed during DNA extraction. In the present study, we assessed the effect of different STHs egg destruction methods, as well as concentration and washing procedures for fecal samples, on the PCR test results. We have shown that bead beating is the most effective and sufficient method for the complete destruction of helminth eggs. In addition, we have shown that parasite concentrators significantly increase the sensitivity of the PCR-based test.

## INTRODUCTION

Soil-transmitted helminths (STHs) infect approximately one billion people worldwide. STHs infection can lead to health problems such as chronic blood loss leading to anemia, malabsorption of nutrients, loss of appetite, lethargy, growth retardation in children, and cognitive decline [1–3]. Currently, the main methods for detecting STHs are microscopic methods [4–6]. With advantages such as a relatively low cost and no need for expensive and difficult-to-maintain equipment, these methods are also quite subjective and require high qualification of the performer, which leads to frequent false negative results [7–10]. Thus, there is a need to develop more sensitive, less time-consuming and high-throughput methods for detecting STHs.

Recently, an increasing number of works have appeared on the use of molecular methods for the diagnosis of STHs. In most studies comparing the sensitivity of methods using microscopy and molecular diagnostic methods, the latter had higher sensitivity [11]. In addition, molecular methods are very specific compared to microscopy and make it possible to distinguish between morphologically related species [8,12]. Molecular methods also make it possible to detect changes in the genome associated with resistance to anthelmintics [13,14].

At the same time, the development of highly sensitive methods for molecular diagnostics of helminthiases requires solving a number of complex technical problems. First, the shell of helminth eggs is quite strong, and it is necessary to select sufficiently harsh conditions to disrupt it while not damaging the helminth DNA [11]. The second problem is the complex composition of the tested clinical material. Feces is a multicomponent matrix containing a variety of compounds, including bile acids and other substances that inhibit amplification reactions [15]. To solve these problems, various research groups have proposed either proprietary protocols [16,17] or used commercially available nucleic acid isolation kits modified with additional processing steps for clinical material, such as adding sorbents, freeze‒thaw, heating, mechanical processing with beads, treatment with enzymes, etc. [18–20]. At the same time, the effectiveness of most of the additional processing steps used is not completely clear, and the lack of generally recognized methods for isolating STHs DNA leads to large scatter in the assessment of the sensitivity of molecular methods for diagnosing diseases caused by these parasites [11].

In this regard, the objectives of this study were, first, to evaluate the effect of the most common methods of destroying STHs eggs on the efficiency of DNA extraction from *T. canis* eggs, both using PCR and visually, and to calculate the concentration of the remaining undamaged eggs in the studied samples using a microscope. Second, we developed an efficient protocol for STHs DNA extraction using commercially available solutions.

## MATERIALS AND METHODS

### Preparation of *T. canis* egg suspension

A suspension of Toxocara eggs in saline was provided by the Laboratory of Microbiology and Parasitology of the Federal State Budgetary Institution “Centre for Strategic Planning and Management of Biomedical Health Risks” of the Federal Medical Biological Agency (Moscow, Russia). The Toxocara eggs were concentrated from the feces of infected dogs and then resuspended in saline (0.9% NaCl). The concentration of the eggs in the suspension was assessed visually using a microscope.

The concentration of the Toxocara eggs was adjusted to 1500 eggs/ml. If the concentration of eggs was greater than this value, the suspension was diluted to the desired concentration with saline. If the concentration of eggs in the suspension was less than 1500 eggs/ml, the suspension was spun at 10000 × g for 5 minutes to increase the concentration, after which the required amount of supernatant was taken, and then the sediment containing the eggs was resuspended.

#### Measurement of Toxocara canis egg concentration by microscopy

The concentration of eggs in the suspension was assessed visually using a Micromed 1 microscope (Micromed, Russia). Ten microliters of the suspension was applied to a glass microscope slide, after which it was covered with a cover slip. The entire preparation was viewed under a microscope using 40X magnification, and the number of eggs in the preparation was counted. This procedure was repeated 10 times, after which the arithmetic average of the number of eggs for 10 specimens was calculated, and the result was multiplied by 100 to obtain the concentration of eggs per milliliter.

### Obtaining model samples of feces contaminated with *T. canis* eggs

Fecal samples (n=52) from patients without signs of helminth infection were obtained from the G.N. Speransky Children’s City Clinical Hospital No 9 (Moscow, Russia). The amount of feces in each sample was at least 7 g.

The Toxocara egg shell has a similar composition and type of structure to egg shells of other STHs found in human feces [21,22], which allows extrapolation of the results of this study to other STH species whose eggs are found in human feces. At the same time, since *T. canis* does not reach the sexually mature stage in the human body and, therefore, cannot produce eggs in the intestine [23], the use of Toxocara eggs as a model object completely excluded their presence in fecal samples from humans before the contamination procedure and made it possible to exactly provide the declared concentration of eggs in different fecal samples.

Fecal samples contaminated with *T. canis* eggs at concentrations of 1000, 500, 50, 10 and 5 eggs per gram of feces were prepared by adding the appropriate amount of Toxocara egg suspension to the sample. In experiments on the effect of preliminary washings and the concentration of helminth eggs on the results of PCR after the isolation of *T. canis* DNA from native feces, fecal samples with a concentration of 1000 eggs/g were used. In the experiment to evaluate the minimum detectable number of *T. canis* eggs in native feces, fecal samples with concentrations of 1000, 500, 50, 10 and 5 eggs/g were used.

### Comparison of the effect of various destructive methods on the integrity of *T. canis* eggs and the efficiency of DNA extraction

To assess the effect of various methods on the integrity of eggs and the efficiency of *T. canis* DNA extraction, we selected four destructive methods: freeze-heat cycles, bead beating, the CD1 lysing buffer of the QIAamp PowerFecal Pro kit, and temperature-dependent enzymes from the forensicGEM Sperm kit. The shells of SHT eggs are largely composed of protein; therefore, proteinases are often used to destroy them [14,24]. For this reason, we decided to test temperature-dependent proteinases from the forensicGEM Sperm kit [25], which are potentially capable of effectively destroying the walls of STH eggs.

Egg integrity was assessed using a light microscope by counting the number of remaining undamaged eggs in the suspension. For the experiment, five 50 µl aliquots of Toxocarа egg suspension (1500 eggs/ml) were taken, which were diluted with saline to a final volume of 100 µl. Tubes with aliquots of Toxocara egg suspension were labeled according to the exposure method: Susp - pure suspension (control); CD1 - incubation in CD1 buffer of the QIAamp PowerFecal Pro kit; Frz-ht - four cycles of freezing-heating; Enzs, temperature-dependent enzymes from the forensicGEM Sperm kit; BB - bead beating with QIAamp PowerFecal Pro ceramic beads.

Evaluation of the destruction methods of Toxocara eggs for their ability to increase the yield of DNA was carried out using PCR. For each exposure method, 134 μl of Toxocara egg suspension (1500 eggs/ml, 200 eggs per sample) was taken in 10 repeats. The names of the experimental groups were similar to the names of the test tubes for assessing the integrity of the eggs, except for the Susp group, which was absent from this experiment. After the initial disruption, DNA extraction from the Toxocara egg suspension was performed using the QIAamp PowerFecal Pro kit. The volume of DNA elution for all samples was 100 µl.

The sample preparation protocols are given in Table 1.

**Table 1.** Sample preparation protocols for evaluating the effect of various destructive methods on the integrity of eggs and the efficiency of Toxocara canis DNA extraction

| Sample | Protocols for Assessing Egg Integrity | Protocols for evaluating the efficiency of DNA extraction |
| --- | --- | --- |
| Susp | 1. Sample was centrifuged with 100 µl of *Toxocara* egg suspension (750 eggs/ml) at 10,000 g, 5 minutes;<br>2. Without discarding the supernatant, resuspend the sediment. | The study was not carried out |
| CD1 | 1. To 100 µl of *Toxocara* egg suspension (750 eggs/ml) 400 µl of CD1 buffer from the QIAamp PowerFecal Pro kit was added;<br>2. Incubated in a thermoshaker for 30 minutes, at a speed of 300 rpm at 25°C;<br>3. The sample was centrifuged at 10,000 × g for 5 minutes, 400 µl of the supernatant was taken, after which the sediment was resuspended in the remaining liquid. | 1. Added to ten 1.5 ml tubes, 134 µl of *Toxocara* egg suspension (1500 eggs/ml), added 800 µl of CD1 buffer from the QIAamp PowerFecal Pro kit;<br>2. The tubes were incubated in a thermoshaker for 30 minutes at a speed of 300 rpm at 25 °C;<br>3. Further DNA extraction was carried out according to the instructions for the QIAamp PowerFecal Pro kit [26], starting from step 4 (excluding the homogenization stage), the entire content was transferred into 2 ml tubes. |
| Frz-ht | 1. The sample was incubated with 100 µl of *Toxocara* egg suspension (750 eggs/ml) in a freezer at -70 °C for 30 min;<br>2. The sample was incubated in a thermoshaker at a speed of 300 rpm at 95 °C for 10 min;<br>Repeat steps 1 and 2 four times; | 1. Added to ten 1.5 ml test tubes, 134 µl of *Toxocara* egg suspension (1500 eggs/ml);<br>2. Samples were incubated in a freezer at -70°C for 30 min;<br>3. Samples were incubated in a thermoshaker at a speed of 300 rpm at 95°C for 10 min; |
|  | 3. The sample was centrifuged at $10,000 \times g$ for 5 minutes, the sediment was resuspended without discarding the supernatant. | Repeat steps 2 and 3 four times;<br>3. Added 800 μl of the QIAamp PowerFecal Pro kit to the samples, incubated for 5 minutes at room temperature;<br>4. Further DNA extraction was carried out according to the instructions for the QIAamp PowerFecal Pro kit [26], starting from step 4 (excluding the homogenization stage), all of the contents were transferred into 2 ml tubes. |
| Enzs | 1. A sample was centrifuged with 100 μl of a suspension of Toxocara eggs (750 eggs/ml) at $10,000 \times g$, 5 minutes, 90 μl of the supernatant was taken, after which the sediment was resuspended in the remaining liquid;<br>2. Added reagents from the forensicGEM Sperm kit: 10 μl buffer, 10 μl Acrosolv, 2 μl forensicGEM, 68 μl deionized water;<br>3. Samples were incubated in a thermoshaker at a speed of 300 rpm for 30 min at 52°, then 30 min at 75°, then 10 min at 95° | 1. Added to ten 1.5 ml tubes 134 μl of Toxocara egg suspension (1500 eggs/ml);<br>2. The tube was centrifuged at $10,000 \times g$ for 5 minutes, 124 μl of the supernatant was taken, after which the sediment was resuspended;<br>3. Added reagents from the forensicGEM Sperm kit: 10 μl buffer, 10 μl Acrosolv, 2 μl forensicGEM, 68 μl deionized water;<br>4. Samples were incubated in a thermoshaker at 300 rpm for 30 min at 52°, then 30 min at 75°, then 10 min at 95°;<br>5. 800 μl of CD1 buffer from the QIAamp PowerFecal Pro kit was added to the samples and incubated for 5 minutes at room temperature;<br>6. Further DNA extraction was carried out according to the instructions for the QIAamp PowerFecal Pro kit [26], starting from step 4 (excluding the homogenization stage), all contents were transferred into 2 ml tubes. |
| BB | 1. Transfer 100 µl of Toxocara egg suspension (750 eggs/ml) to the beating tube from the QIAamp PowerFecal Pro kit, add 800 µl of saline;<br>2. Homogenize the sample on a QIAamp TissueLyser homogenizer at 50 Hz for 30 minutes;<br>3. Immediately after homogenization, 600 µl of the upper part of the homogenate was taken, without capturing the beads, and transferred to a clean 1.5 ml tube;<br>4. The tube was centrifuged at $10,000 \times g$ for 5 minutes, 500 µl of the supernatant was taken, after which the sediment was resuspended;<br>5. Added 600 µl of saline to the same beating tube, vortexed for 5 seconds at maximum speed, after which 600 µl of liquid was taken into the tube with the homogenate sediment;<br>6. The tube was centrifuged at $10,000 \times g$ for 5 minutes, 600 µl of the supernatant was taken, after which the sediment was resuspended in the remaining liquid;<br>Repeat steps 5 and 6 four times | 1. Added to ten PowerBead Pro Tubes from the QIAamp PowerFecal Pro kit 134 µl of Toxocara egg suspension (1500 eggs/ml);<br>2. 800 µl of CD1 buffer from the QIAamp PowerFecal Pro kit was added to the samples, mixed on a vortex;<br>3. Homogenize the samples on a QIAamp TissueLyser homogenizer at 50 Hz for 30 minutes;<br>4. Further DNA extraction was carried out according to the instructions for the QIAamp PowerFecal Pro kit [26], starting from step 3 (centrifugation step after homogenization). |

As seen from the above protocols for preparing samples for microscopic examination, the final volume of the resuspended sediments of the homogenates after all manipulations was the same for all aliquots at 100 µl. Since the CD1, Enzs and BB samples had to be centrifuged to remove the excess fluid volume, the Susp and Frz-ht samples were subjected to the same centrifugation procedure to take into account the possible effect of centrifugation on the integrity of the helminth eggs. The number of undamaged eggs in the resulting sediments of the homogenates was assessed under a microscope, as described above. For each sample, 8 specimens were made, after which the counting data for each specimen were analyzed.

#### qPCR

Primers for the *T. canis* internal transcribed spacer 1 (ITS1) conserved region were selected using the NCBI Primer Blast web service [27] based on the ITS1 nucleotide sequences of the *T. cati* and *T. canis* species available in the NCBI Nucleotide Database [28]. The resulting primer pairs, together with the nucleotide sequences of Toxocara and related roundworm species, were aligned using the Unipro Ugene software, version 41.0, using the Clustal W algorithm. TaqMan probes [29,30]. The thermodynamic characteristics of the primers, the fluorescent probe, and their secondary structures were evaluated using the Themfold Web Server online service [31]. Synthesis of the primers and probes was carried out by JSC Genterra.

The primer and probe sequences are shown in Table 2.

**Table 2.**
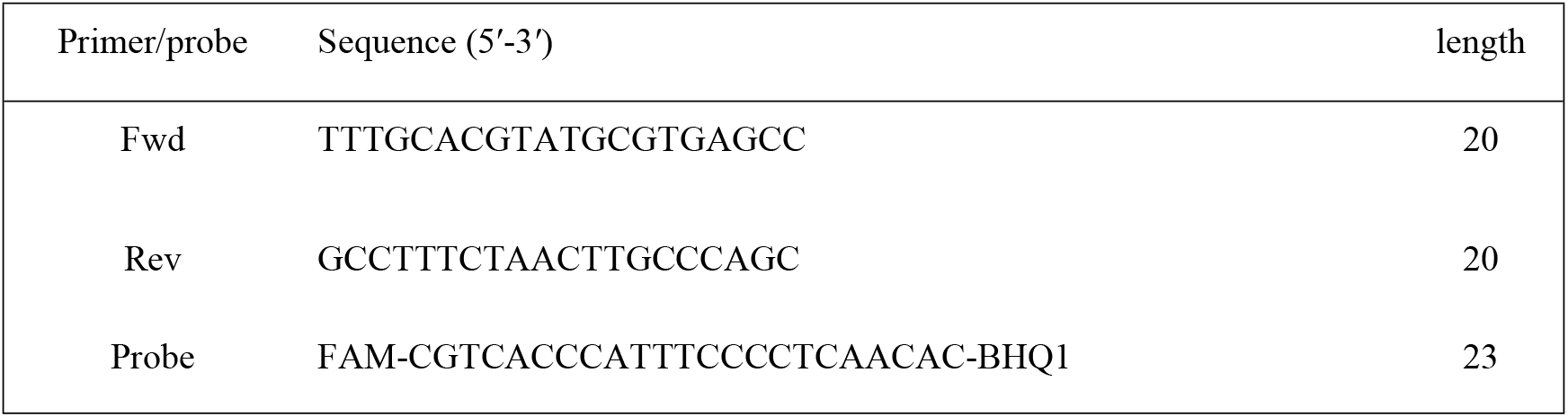
Primers and probe for *T. canis* detection

The reaction mixture with a total volume of 25 µl contained the following components: 10 µl of extracted DNA solution, 3.2 nM of each primer, 2.0 nM probe and 5 µl of 5x Genta qPCR MasterMix (JSC Genterra, Russia). PCR was performed on a CFX96 RealTime PCR amplification platform (Bio-Rad, USA).

The amplification program included the following stages of thermal cycling: 95 °C for 15 min, followed by 45 cycles of 95 °C for 15 s, 58 °C for 30 s, and 72 °C for 15 s. The fluorescence accumulation signal of the target DNA was recorded using the channel for the FAM fluorophore.

The result was evaluated by the threshold method, determining the threshold cycles of Ct amplification by the intersection of the fluorescence curve with the threshold line set in the middle of the exponential section of the fluorescence increase graph on a logarithmic scale.

### Study of the influence of preliminary washings and concentration of helminth eggs on the results of PCR for the extraction of *Toxocara canis* DNA from native feces

#### DNA extraction

For the experiment on the effect of preliminary washings on the PCR results from the extraction of *T. canis* DNA from native feces, aliquots of fecal samples contaminated with Toxocara eggs at a concentration of 1000 eggs/g were preliminarily washed twice with 0.1% Tween-20 (Merck Life Science, Germany) diluted in phosphate buffered saline (VWR international LCC, USA). These samples constituted experimental Group W. The reference groups were fecal samples contaminated with Toxocara eggs without prewash (Fec), as well as samples of a pure suspension of Toxocara eggs (Pure). The DNA extraction protocols for the experimental groups are shown in Table 3. The DNA elution volume for all samples was 100 µl.

**Table 3.** DNA extraction protocols for the experiment on the effect of prewashes on PCR results

| Group | Protocol |
| --- | --- |
| Pure (n=10) | 1. Ten PowerBead Pro Tubes from the QIAamp PowerFecal Pro kit had 134 µl of *Toxocara* egg suspension added;<br>2. 800 µl of CD1 buffer from the QIAamp PowerFecal Pro kit was added to the samples, mixed on a vortex;<br>3. Homogenize the samples on a QIAamp TissueLyser homogenizer at 50 Hz for 30 minutes;<br>4. Further stages of DNA extraction were carried out according to the instructions for the QIAamp PowerFecal Pro kit [26], starting from step 3. |
| Fec (n=10) | 1. Ten PowerBead Pro Tubes from the QIAamp PowerFecal Pro kit were filled with 200 mg of feces contaminated with *Toxocara* eggs at a concentration of 1000 eggs/g; |
|  | 2. 800 µl of CD1 buffer from the QIAamp PowerFecal Pro kit was added to the samples, mixed on a vortex;<br>3. Homogenize the samples on a QIAamp TissueLyser homogenizer at 50 Hz for 30 minutes;<br>4. Further stages of DNA extraction were carried out according to the instructions for the QIAamp PowerFecal Pro kit [26], starting from step 3 (the stage of sample centrifugation after homogenization). |
| W (n=10) | 1. Ten 2 ml test tubes were filled with 200 mg of feces contaminated with *Toxocara* eggs at a concentration of 1000 eggs/g;<br>2. 1.5 ml of a 0.1% solution of Tween-20 in PBS was added to the tubes, after which the tubes were vortexed until the feces were completely dissolved;<br>3. The tubes were centrifuged for 5 min at 5000 g, after which the supernatant was merged;<br>Steps 2 and 3 were repeated 2 times<br>4. The sediment was resuspended in 800 µl of CD1 buffer from the QIAamp PowerFecal Pro kit, vortexed, and then the contents of the tubes were transferred to clean PowerBead Pro Tubes from the QIAamp PowerFecal Pro kit;<br>5. Homogenize the samples on a QIAamp TissueLyser homogenizer at 50 Hz for 30 minutes;<br>6. Further stages of DNA extraction were carried out according to the instructions for the QIAamp PowerFecal Pro kit [26], starting from step 3 (centrifugation of the samples after homogenization). |

For the experiment on the effect of concentrating helminth eggs on the PCR results during the extraction of *Toxocara canis* DNA from native feces, aliquots of fecal samples contaminated with Toxocara eggs at a concentration of 1000 eggs/g were preconcentrated using Apacor Mini Parasep concentrators (Apacor, UK), after which they were concentrated once, then washed with a 0.1% solution of Tween-20 in phosphate-buffered saline to wash any formalin out of the obtained concentrates. These samples constituted the W+PS experimental group. The reference groups were samples of feces contaminated with Toxocara eggs, previously washed twice with Tween-20 (W) solution, as well as samples of a pure suspension of Toxocara eggs (Pure). The protocols for DNA extraction for the Pure and W experimental groups correspond to the protocols given in Table 3. For the W+PS group, the following protocol was used:

1. We added 500 mg of feces contaminated with Toxocara eggs at a concentration of 1000 eggs/g to the sample collection unit of the Apacor Mini Parasep concentrators.
2. The samples were mixed and centrifuged in accordance with the instructions for the concentrators (EU Protocol v3.0 2017.09) [32].
3. A total of 1.5 ml of a 0.1% solution of Tween-20 in PBS was added to the sediments, after which the tubes were vortexed until the feces were completely dissolved.
4. The tubes were centrifuged for 5 min at 5000 g, after which the supernatant was merged.
5. The sediment was resuspended in 800 µl of CD1 buffer from the QIAamp PowerFecal Pro kit, vortexed, and then the contents of the concentrator were transferred to clean PowerBead Pro Tubes from the QIAamp PowerFecal Pro kit.
6. Further stages of DNA extraction were carried out according to the instructions for the QIAamp PowerFecal Pro kit [26], starting from step 3 (the stage of sample centrifugation after homogenization).

The volume of DNA elution for all samples was 100 µl.

#### qPCR

Real-time PCR was performed as described above. In these experiments, together with the threshold amplification cycles Сt, the maximum fluorescence intensity (ΔRFU) was determined, which was calculated for each sample as the difference between the fluorescence intensity at the end of the PCR (on the 45th amplification cycle) and at the beginning (on the 10th amplification cycle), when the signal fluorescence had stabilized, and the exponential growth phase of fluorescence had not yet begun.

### Statistical data analysis

Statistical data analysis was performed using the Statistica 12 software package (TIBCO Software Inc., USA). We used the functions of the “Nonparametric statistics” section. The “Mann‒Whitney U Test” function was used to evaluate the differences between the two groups. To assess the differences between three or more groups, the functions “Summary: Kruskal‒Wallis ANOVA & median test” and “Multiple comparisons of mean ranks for all groups” were used, where the dependent variables were Ct or ΔRFU values, and the experimental groups were independent variables. Comparisons of the PCR parameters were carried out within one setting. Differences with a significance level of p≤0.05 were recognized as statistically significant.

## RESULTS

### Comparison of methods for destroying *T. canis* eggs using microscopy

Exposure of Toxocara egg suspension to freeze-heat cycles, temperature-dependent enzymes from the forensicGEM Sperm kit, and incubation with CD1 lysis buffer from the QIAamp PowerFecal Pro kit did not lead to a significant change in the number of undamaged eggs in the suspension. However, after bead beating with beads from the QIAamp PowerFecal Pro kit, no undamaged eggs were found in the samples, while several objects were found that were presumably broken eggs (Fig. 1, Fig. 2).

**Figure 1.**
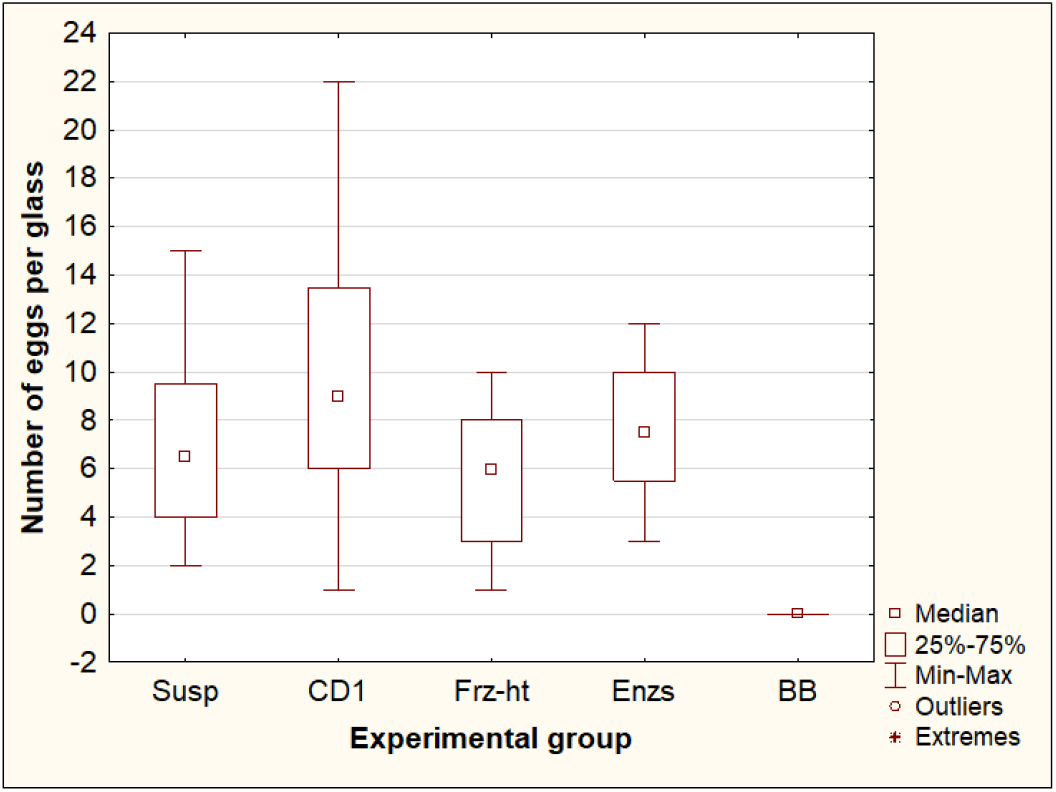
Comparison of the number of whole eggs of *T. canis* after processing by different methods. Susp – initial suspension of eggs (control, n=8); **CD1** – incubation in CD1 buffer from the QIAamp PowerFecal Pro kit (n=8); **Frz-ht** – freeze-heat cycles (n=8); **Enzs** – incubation with temperature-dependent enzymes from the forensicGEM Sperm kit (n=8); **BB** - bead beating with beads from the QIAamp PowerFecal Pro kit (n=8).

**Figure 2.**
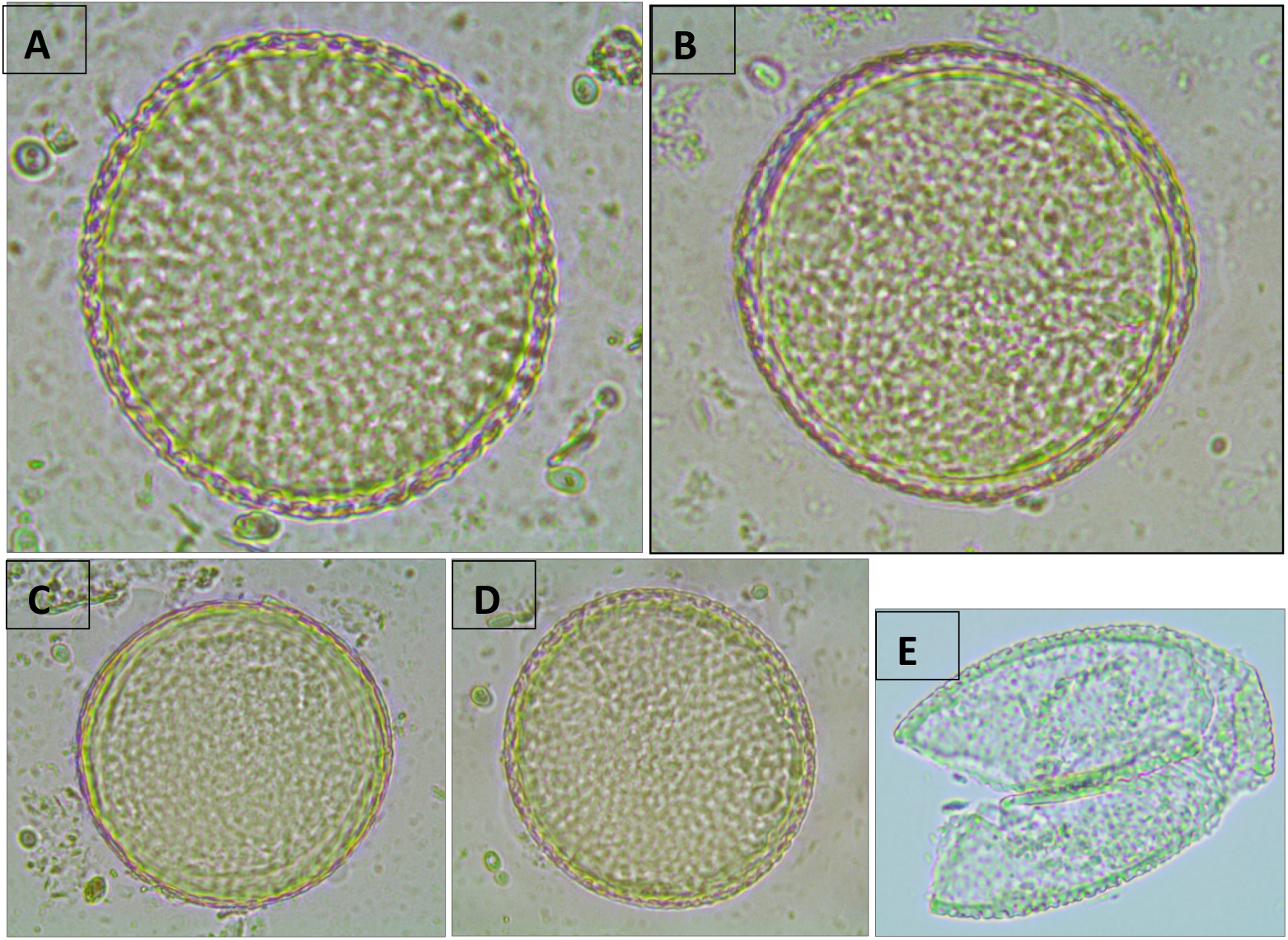
Representative photos of *Toxocara canis* eggs after treatment with various methods (400X magnification). **A**, Initial suspension of eggs; **B**, incubation in CD1 buffer from the QIAamp PowerFecal Pro kit; **C**, freeze-heat cycles; **D**, treatment with temperature-dependent enzymes from the forensicGEM Sperm kit; **E**, bead beating with beads from the QIAamp PowerFecal Pro kit.

### Comparison of the yield of DNA from *T. canis* eggs by PCR as a result of the action of various destructive methods

Of the four methods for destroying Toxocara eggs, only bead beating and incubation in CD1 buffer from the QIAamp PowerFecal Pro kit resulted in the release of Toxocara DNA in all repeats (10 out of 10 for each experimental group). When comparing these two methods (Figure 3), bead beating resulted in a significant decrease in Ct values (p=0.0005). The difference between the median Ct values was 2.7, which suggests that bead beating increased the yield of DNA from Toxocara eggs by approximately 6.4 times. Two other methods, freeze-heat cycles and incubation of Toxocara eggs with temperature-dependent enzymes, only sporadically resulted in DNA release at low concentrations. After freeze-heat cycles, a positive PCR result was obtained for one sample out of 10 (Ct=44.4). After incubation with enzymes from the forensicGEM Sperm kit, only 2 out of 10 samples gave a positive PCR result (Ct=38.5; 40.4).

**Figure 3.**
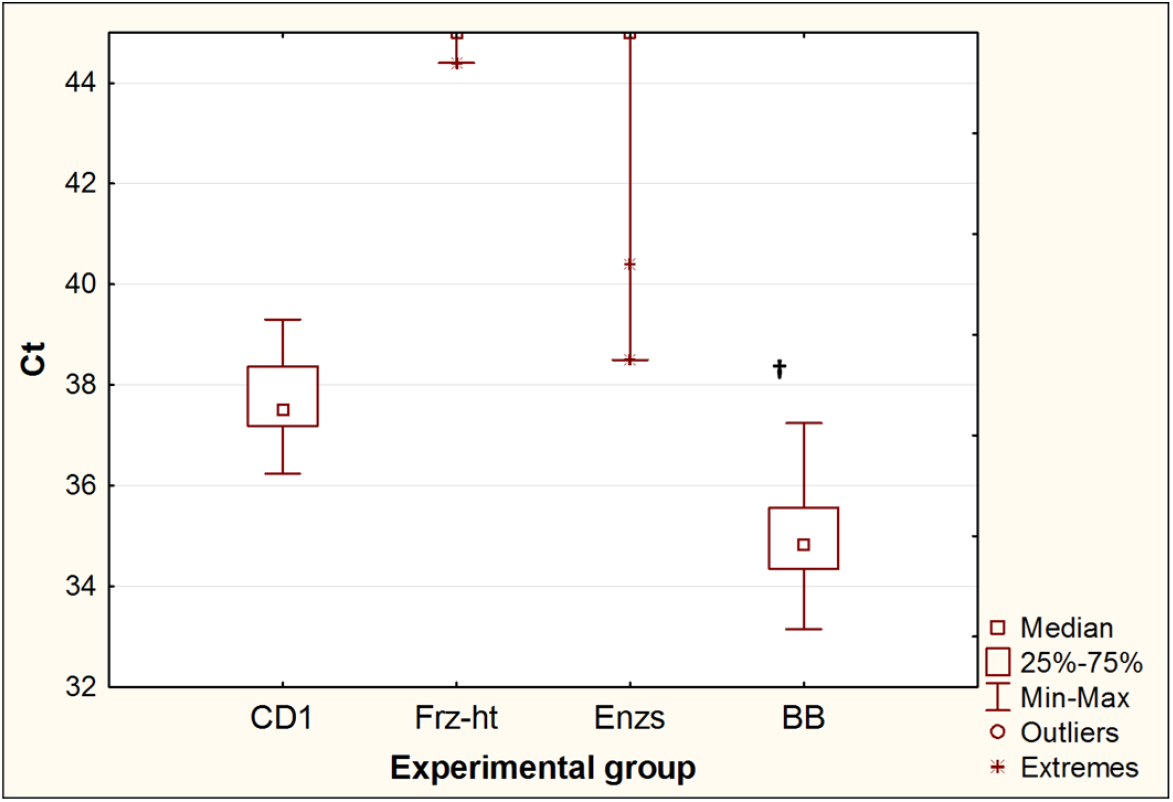
Comparison of threshold amplification cycles (Ct) of DNA after egg disruption by different methods. **CD1** – incubation in CD1 buffer from the QIAamp PowerFecal Pro kit (n=10); **Frz-ht** – freeze-heat cycles (n=10); **Enzs** – treatment with temperature-dependent enzymes from the forensicGEM Sperm kit (n=10); **BB** - bead beating with beads from the QIAamp PowerFecal Pro kit (n=10); **†** – p<0.05 vs. CD1.

### Effect of double prewash on the PCR results in the extraction of *T. canis* DNA from native feces

The extraction of *T. canis* DNA from contaminated feces using the QIAamp PowerFecal Pro kit for PCR showed an increase in Ct (p= 0.015) and a decrease in the maximum fluorescence intensity ΔRFU (p= 0.019) compared with DNA isolated with the same kit from the same number of eggs in the pure Toxocarа egg suspension. The difference between the median Ct values for DNA isolated from feces and DNA isolated from a pure Toxocara egg suspension was 2.7, which implies that when DNA is isolated from feces, without their preliminary purification from inhibitors, the amount of *T. canis* DNA detected by PCR decreases approximately 6.4 times. Preliminary double washing of feces contaminated with *T. canis* eggs with a 0.1% solution of Tween-20 in PBS significantly reduced the Ct values (p= 0.0013) and increased the ΔRFU (p= 0.0082), bringing these values closer to the values obtained by isolating DNA from a pure suspension of Toxocara eggs (Fig. 4).

**Figure 4.**
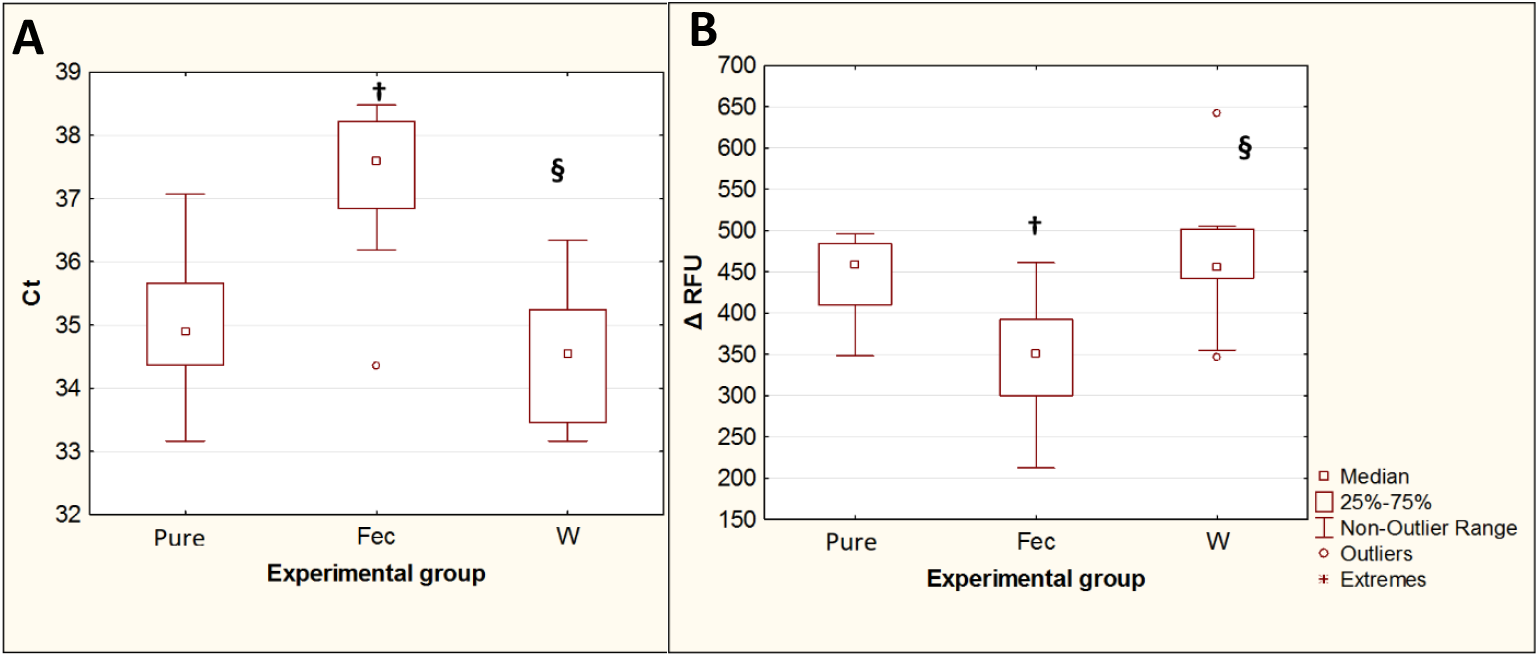
Comparison of PCR results for isolation of *T. canis* DNA from native feces and from feces after double prewashing. **A** - threshold amplification cycles (Ct). **B** - maximum fluorescence intensity (ΔRFU). **Pure** - pure suspension of Toxocara eggs (n=10); **Fec** – feces contaminated with Toxocara eggs (n=10); **W** – feces contaminated with Toxocara eggs after double preliminary washing (n=10). **†** – p<0.05 vs. Susp; **§** - p<0.05 vs. Fec.

### Influence of preconcentration of helminth eggs on PCR results in isolation of *T. canis* DNA from native feces

Replacing the first of the two prewashes with the Apacor Parasep mini concentrator significantly reduced the Ct fluorescence threshold cycles (p= 0.0016) and increased the maximum fluorescence intensity ΔRFU (p= 0.00007) compared to the isolation of Toxocara DNA from contaminated feces and using a double prewash (Figure 5). The difference between the median Ct values for these two experimental groups was 4, which implies that when DNA is isolated from feces, after preliminary concentration, the amount of *Toxocara canis* DNA detected by PCR increases by approximately 16 times.

**Figure 5.**
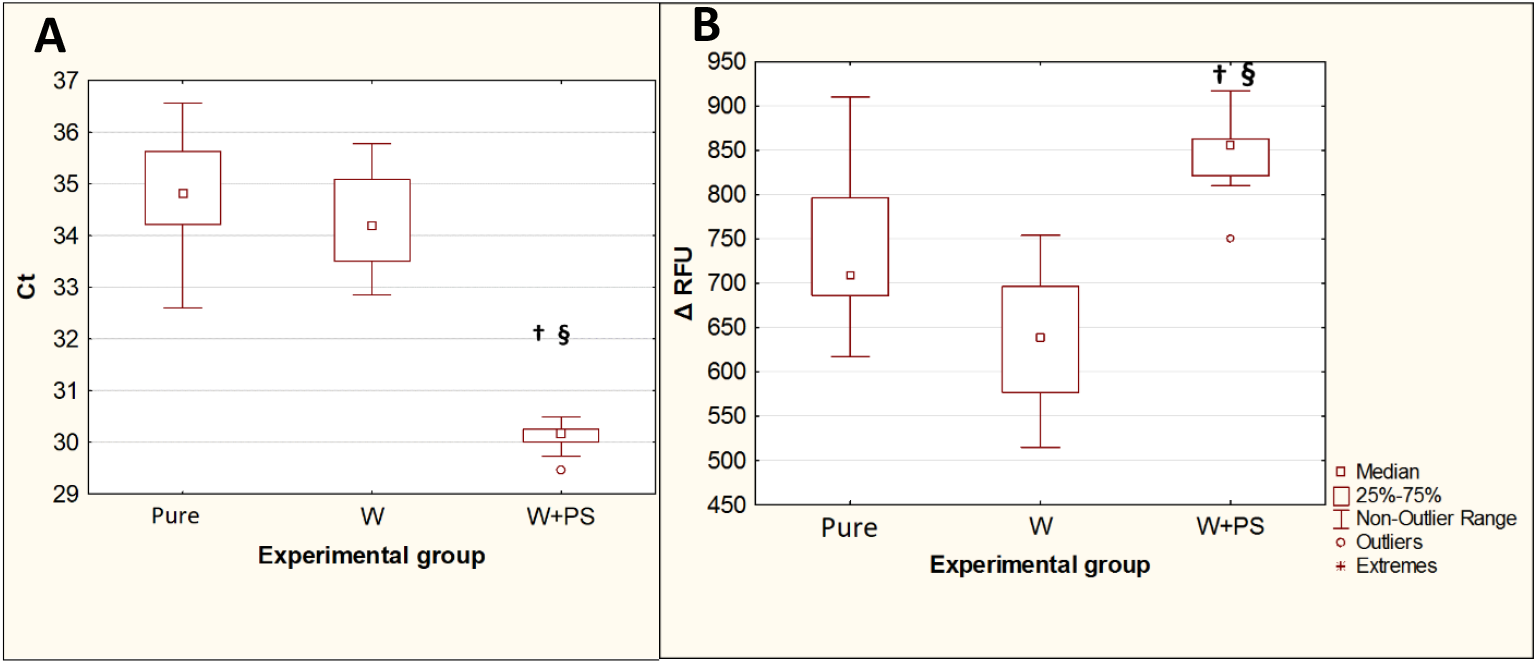
Comparison of the PCR results for isolation of *T. canis* DNA from feces after a double prewash and using a concentrator followed by a single wash. **A** - threshold amplification cycles (Ct). **B** - maximum fluorescence intensity (ΔRFU). **Pure** - pure Toxocara egg suspension (n=10); **W** – feces contaminated with Toxocara eggs after preliminary double washing (n=10); **W+PS** – feces contaminated with Toxocara eggs after preconcentration followed by single washing (n=10). **†** – p<0.05 vs. Susp; **§** - p<0.05 vs. W.

### Evaluation of the minimum detectable number of eggs of *Toxocara canis* in native feces by real-time PCR using the developed protocol for the isolation of STHs DNA from feces

This experiment used fecal samples contaminated with *T. canis* eggs at concentrations of 1000, 500, 50, 10 and 5 eggs/g, each in 10 repeats.

The use of the W+PS protocol with a preliminary concentration of helminth eggs with an Apacor Parasep mini concentrator, washing with 0.1% Tween-20 solution in PBS, and further DNA isolation from feces using the QIAamp PowerFecal Pro kit, made it possible to reproducibly detect Toxocara DNA in all repeats at concentrations of Toxocara eggs of up to 50 eggs per gram of feces. At a concentration of 10 eggs per gram of feces, Toxocara DNA was detected in 40% of samples (4 out of 10). A concentration of 5 eggs per gram of stool was insufficient to detect *T. canis* DNA by real-time PCR (Figure 6).

**Figure 6.**
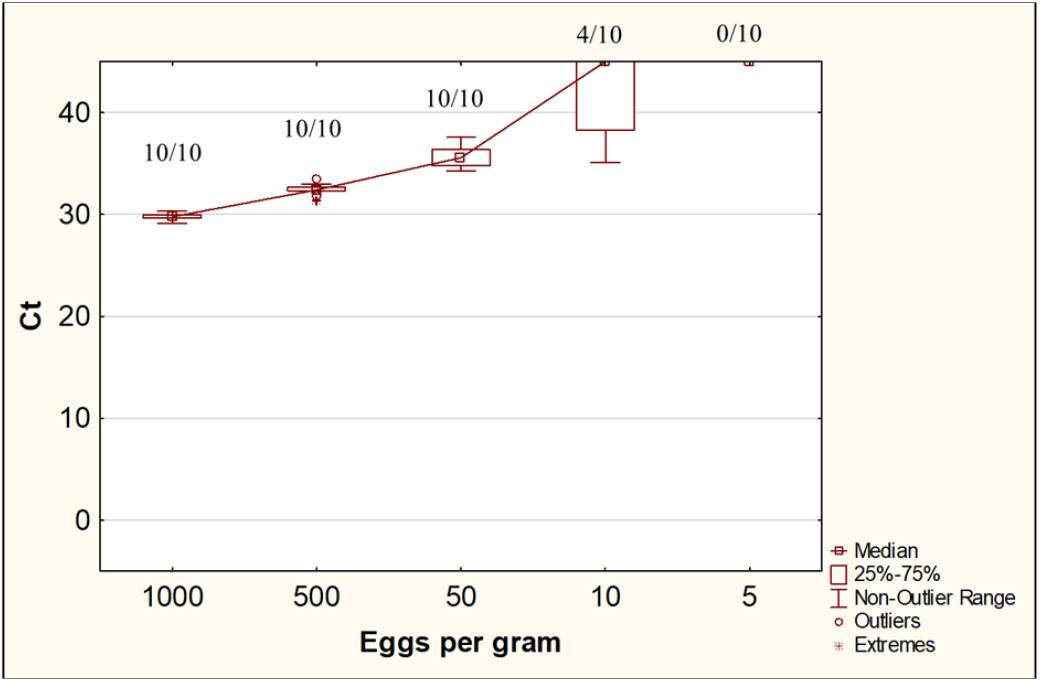
Ct values for DNA isolation from feces contaminated with *Toxocara canis* eggs at various concentrations. The numbers at the top of the graph are the ratio of positive samples according to the results of the PCR study to the total number of samples in the group.

## DISCUSSION

Experiments comparing different methods of destroying Toxocara eggs showed that bead beating significantly increased the yield of DNA from the eggs. These data are consistent with the results of previous studies both on helminths of the genus Toxocara [33] and on other STHs [34,35]. At the same time, such a widely used destruction method as exposing helminth eggs to high and low temperatures [13,36–38] did not lead to a significant increase in the number of destroyed eggs observed by microscopy or to a significant release of *T. canis* DNA from the eggs. Similarly, the effectiveness of temperature-dependent enzymes on the destruction of eggs and the release of helminth DNA has not been shown. In protocols that use enzymatic treatment of helminth eggs, incubation with the enzymes takes from several hours to almost a day [13,24,39]. This far exceeds the time required for complete destruction of STH eggs by the bead beating method demonstrated in this study.

Unexpectedly, there was a discrepancy between the results on the action of the CD1 lysis buffer from the QIAamp PowerFecal Pro kit, obtained by microscopy (the number of undamaged eggs did not change significantly) and by real-time PCR (*T. canis* DNA was detected). The presence of *T. canis* DNA in the samples with a constant number of undamaged eggs can be explained by two reasons. First, in addition to eggs, fragments of helminth tissues could be present in the suspension. In this case, the CD1 buffer could lyse these particles and release their DNA, leaving the eggs intact. The second explanation may be that Toxocara eggs, when incubated in CD1 buffer, undergo damage that is not visible under a light microscope but is sufficient to release a significant amount of DNA into the solution. QIAGEN does not disclose the exact composition of its solutions; however, from the documents for the QIAamp PowerFecal Pro kit published on the company’s website [40], it can be concluded that the main lysing component of the CD1 solution is the relatively mild chaotropic agent sodium thiocyanate. According to patent CA3096461A1 from Qiagen Sciences LLC, dedicated to the isolation of nucleic acids and the removal of inhibitors from complex samples, the use of mild lysing chemical agents makes it possible to reduce the loss of isolated nucleic acids but requires additional physical treatment of the sample, for example, by bead beating [41]. However, short-term incubation in a CD1 solution for 5 minutes, in which the samples were subjected to freeze-heat cycles and after the action of temperature-dependent enzymes, did not lead to a significant yield of DNA. Damage to Toxocara eggs can be detected, for example, by immunohistochemistry methods, but we did not use these methods, since this was not the main purpose of this study. However, based on the data obtained, we can conclude that the bead beating procedure may be sufficient for the complete destruction of STH eggs, while additional steps for egg destruction, such as temperature or enzyme action, are not necessary.

At the same time, data on the effect of preliminary washings of feces with 0.1% Tween-20 solution in PBS show that when using this kit for the isolation of STHs DNA from feces, it is desirable to use additional steps aimed at removing PCR inhibitors. Despite the fact that in the experiment on the effect of preliminary washings of feces in all experimental groups the number of Toxocara eggs was approximately the same (approximately 200 eggs per sample), the amount of *T. canis* DNA detected when isolated from feces was several times lower than when isolated from pure egg suspension. A significant decrease in ΔRFU was also noted when DNA was isolated from contaminated feces compared with a pure Toxocara egg suspension. ΔRFU depends on many parameters of the reaction mixture, including the fluorescent label used, the ionic strength of the solution, the cycler parameters, etc. At the same time, a decrease in ΔRFU when using the same reagents within one setting on the device may indicate the presence of PCR inhibitors in the sample [42,43]. At the same time, preliminary washings of fecal samples with a simple solution of a nonionic surfactant in a buffer can significantly reduce PCR inhibition, which is expressed by a decrease in Ct and an increase in ΔRFU to the levels achieved when Toxocara eggs are isolated from a pure suspension of eggs. Thus, prewashing of fecal samples before the isolation of parasitic DNA is an important step that can significantly improve the PCR results.

The concentration of STHs eggs prior to DNA isolation can significantly increase the sensitivity of molecular diagnostic methods for helminth infections in low-incidence regions, where patients with a low helminth invasion range often have relatively few parasite eggs in their feces. The use of commercially available parasite concentrators for feces is a fairly common practice in parasitological studies using microscopy methods [44–49]. At the same time, we found a small number of works where protocols for isolating helminth DNA from feces used flotation– sedimentation for concentrating parasite eggs [50–52]. At the same time, in most of these studies, the eggs were washed off the coverslips for further DNA isolation after flotation, which greatly complicates the DNA isolation procedure and increases the risk of losing some of the parasite eggs. In addition, these works did not evaluate the effectiveness of the removal of PCR inhibitors by the flotation methods. Studies on the use of commercially available parasite egg concentrators for the isolation of STH DNA from feces are not found in the literature.

Theoretically, in addition to physical methods, it is possible to concentrate STH eggs from feces by immunosorbent methods. The literature describes protocols for isolating DNA of protozoan parasites with concentration using the method of immunomagnetic separation [53]. However, this method is quite laborious and much more expensive than the parasite concentrator, and before its use, a step of preliminary isolation of parasite cysts from feces is still necessary, for example, by flotation. In addition, existing commercially available kits are designed for the immunomagnetic separation of protozoan parasites (*Cryptosporidium spp*., *Giardia lamblia*, etc.). Isolation of STH eggs by this method is likely to be less efficient due to the much larger size of helminth eggs compared to parasitic protozoan cysts.

In the experiment on the effect of concentration of helminth eggs, in the group with the replacement of the first wash with Apacor Parasep Mini concentrators, due to the ability to use a larger volume of feces for DNA extraction, the number of *T. canis* eggs was approximately 500 eggs per sample, which was 2.5 times higher than the number of eggs per sample in the two wash group (approximately 200 eggs per sample). At the same time, the amount of detectable *T. canis* DNA obtained by adding the concentration step to the DNA isolation protocol increased by an average of 16 times. This suggests that the use of concentrators not only allows the number of STHs eggs to increase in the test sample but also additionally reduces the amount of PCR inhibitors in the sample. At the same time, one washing of the precipitate obtained during the concentration with a Tween-20 solution is sufficient to remove the formalin from the precipitate, which is part of the solution used in the concentrator and is a powerful PCR inhibitor [54].

According to the present study, the use of a modified protocol for the QIAamp PowerFecal Pro excretion kit, with preliminary concentration of the helminth eggs with an Apacor Parasep mini concentrator and a single wash of 0.1% solution of Tween-20 in PBS, made it possible to detect *T. canis* DNA in 100% of samples at concentrations up to 50 eggs per gram of feces and in 40% of samples at a concentration of 10 eggs per gram of feces. For *T. canis*, the mean values for eggs in dog feces range from 400 to 1900 eggs per gram, depending on the age of the dogs and the study region [55,56]. For STHs parasitizing the human intestine, according to WHO recommendations, the boundary values for mild intensity of helminthic invasion range from 1000 (for *Trichuris trichiura*) to 4000 (for *Ascaris lumbricoides*) eggs per gram of feces [57]. *T. canis* was a model object in our study, and the efficiency of DNA extraction for other STHs species may differ. Nevertheless, the obtained results, together with the data presented in the literature, suggest that our proposed protocol has sufficient extraction efficiency to detect human intestinal STH DNA in most cases of mild invasion.

We chose the following protocol for the isolation of STH DNA from feces as the optimal protocol:

1. Unscrew the lid from the sample collection unit of the Apacor Mini Parasep Concentrator (do not discard the lid!), add approximately 500 mg of fecal sample (1 spoonful) into the compartment using the spoon at the end of the filter.
2. Follow the “Emulsification” and “Centrifugation” sections of the instructions for the Apacor Mini Parasep concentrator kit.
3. Gently open the device and discard the top chamber of the concentrator along with the filter.
4. Discard the supernatant, add 1.5 ml of a 0.1% solution of Tween-20 in PBS to the sediment, and then close the tube with the lid from the sample collection compartment and resuspend the sediment.
5. Centrifuge the tubes for 5 min at 5000 g and discard the supernatant.
6. Add 800 µl of CD1 buffer from the QIAamp PowerFecal Pro kit to the sediment, and resuspend the sediment on a vortex.
7. Transfer the resulting suspension to a clean PowerBead Pro Tube from the QIAamp PowerFecal Pro kit.
8. Homogenize the samples on the bead beating homogenizer at maximum speed for 30 minutes.
9. Follow the instructions for the QIAamp PowerFecal Pro kit starting at step 3.

## CONCLUSION

Thus, in this study, we showed that the most complete destruction of STHs eggs is achieved by bead beating, and there is no need for additional steps such as exposure to high/low temperatures or enzyme treatment. The use of protocols for extracting DNA from STH eggs that do not include a bead-beating step is not optimal. Steps such as preconcentration of STH eggs from feces using concentrators, as well as washing the feces concentrate with a nonionic surfactant solution, can significantly increase the amount of DNA extracted from the sample. In summary, we propose a protocol for preparing DNA from feces for use in PCR tests for helminth parasite eggs.

## Data Availability

All relevant data is deposited in Open Science Framework (URL: https://osf.io/2ruyt/)

https://osf.io/2ruyt/files/osfstorage

